# Validation of a Paediatric-Optimized Computer-Aided Detection System for Tuberculosis Using Bayesian Latent Class Analysis

**DOI:** 10.64898/2026.05.16.26353382

**Authors:** Victory F. Edem, Schadrac C. Agbla, Esin Nkereuwem, Sheila A. Owusu, Nuredin Mohammed, Abdou K. Sillah, Omolola M. Atalabi, Uzochukwu Egere, Beate Kampmann, Toyin Togun

**Affiliations:** Medical Research Council Unit The Gambia at London School of Hygiene and Tropical Medicine, Fajara, The Gambia; Department of Immunology, College of Medicine, University of Ibadan, Ibadan, Oyo State, Nigeria; Department of Health Data Science, University of Liverpool, Liverpool, UK; Department of Infectious Diseases Epidemiology, London School of Hygiene and Tropical Medicine, London, UK; Faculty of Infectious and Tropical Diseases, London School of Hygiene and Tropical Medicine, London, UK; University for Development Studies, Tamale, Northern Region, Ghana; Department of Radiology, College of Medicine, University of Ibadan, Ibadan, Oyo State, Nigeria; Liverpool School of Tropical Medicine, Liverpool, United Kingdom; Charité Centre for Global Health, Institute of International Health, Berlin, Germany; TB Centre, London School of Hygiene and Tropical Medicine, London, UK

**Author notes:** Corresponding author: Professor Toyin Togun PhD, London School of Hygiene & Tropical Medicine (LSHTM), London, United Kingdom, and Medical Research Council Unit The Gambia at LSHTM, Fajara, The Gambia.

**Keywords:** Diagnostic accuracy, Paediatric tuberculosis, Computer-aided detection, Digital chest X-ray, Children

## Abstract

**Background:** Microbiological confirmation of paediatric pulmonary tuberculosis is frequently unattainable, rendering chest radiography a critical yet underutilised diagnostic tool.

**Methods:** We conducted a retrospective diagnostic accuracy study of the qXR-version 4.2.1 (Qure.ai), a paediatric-optimized computer-aided detection (CAD) algorithm, for pulmonary tuberculosis. Diagnostic performance was assessed against microbiological (MRS) and clinical reference standards (ClRS). Bayesian latent class analysis (LCA) was applied to address the imperfection of both reference standards in children. Performance was quantified using area under the receiver operating characteristic curve (AUROC) and estimates of sensitivity and specificity.

**Results:** We included digital chest radiographs of 932 Gambian children (< 15 years) comprising 80 (9%) children with confirmed tuberculosis, 163 (17%) with unconfirmed tuberculosis, and 689 (74%) classified as unlikely tuberculosis. Against MRS, qXR demonstrated AUROC, sensitivity and specificity of 0.68 (95% CI 0.61–0.75), 54% (95% CI 43–64%), and 82% (95% CI 79–84%), respectively. Against ClRS, the AUROC, sensitivity and specificity were 0.73 (95% CI 0.69–0.77), 41% (95% CI 34–49%), and 87% (95% CI 84–89%), respectively. Bayesian LCA, assuming conditional independence, estimated sensitivity of 79% (95% CrI 65–89%) and specificity of 82% (95% CrI 79–84%). Assuming conditional dependence between qXR and expert radiologist, and between culture and Xpert, estimated sensitivity increased to 89% (95% CrI 71–98%), with specificity remaining at 82% (95% CrI 79–84%).

**Conclusions:** Paediatric-optimized qXR algorithm provides a valuable complementary tool for diagnosis of paediatric pulmonary tuberculosis. Conventional reference standards likely underestimate the true diagnostic performance of CAD systems in children.

## Introduction

Tuberculosis caused an estimated 1.2 million new cases and over 174,000 deaths among children aged under 15 years in 2024 [1]. Despite these alarming morbidity and mortality rates, modelling studies suggest that at least 60% of tuberculosis cases in children remain undiagnosed and untreated [2]. This diagnostic gap reflects the fundamental difficulty of confirming tuberculosis in children: bacteriological confirmation is achievable in fewer than 50% of treated children, even when WHO-recommended rapid molecular diagnostics (mWRDs) are the initial test [3, 4]. Hence, childhood tuberculosis is predominantly diagnosed presumptively, based on the integration of epidemiological, clinical, and radiological findings without microbiological confirmation [5]. In settings with limited paediatric clinical expertise, this diagnostic dilemma contributes to delays in treatment initiation and poor outcomes, including death [6].

Chest radiography occupies a central but underutilised role in the diagnostic workup of childhood tuberculosis. Although it is frequently the only adjunctive tool available for the clinical evaluation of paediatric pulmonary tuberculosis, its interpretation is inherently subjective, expertise-dependent, and inconsistent across settings [7]. Computer-aided detection (CAD) systems, which apply artificial intelligence to automated analysis of digital chest radiographs (dCXRs), have demonstrated clinical utility for tuberculosis screening and triage in individuals aged 15 years and above, and their use in adults has been endorsed by the WHO [8, 9]. Their deployment in settings with dCXR infrastructure but limited experienced human readers could substantially expand tuberculosis screening coverage and improve case detection rates [8]. However, available evidence indicates that CAD algorithms validated in adults perform substantially less well in children [7].

Children represent a diagnostically distinct population. Tuberculosis in this group is characterised by paucibacillary disease, atypical and heterogeneous radiological findings [10], and overlap of symptoms and signs with other common childhood respiratory conditions such as bacterial pneumonia and viral lower respiratory tract infection [11]. Therefore, CAD systems developed without representative training datasets that capture the full spectrum of paediatric radiological presentations, and that account for the normal anatomical variation across the age spectrum from infancy to adolescence, are likely to underperform in children.

In a previous study of 724 Gambian children under 15 years with presumptive tuberculosis, we provided evidence supporting the need to optimise existing CAD algorithms and to develop paediatric-specific CAD systems capable of aiding tuberculosis detection in children [12]. This evidence was corroborated by Palmer and colleagues, who demonstrated that fine-tuning the CAD4TB-version 7 algorithm (Delft Imaging, the Netherlands) on a training dataset of 445 paediatric dCXRs resulted in improved diagnostic accuracy in a test set of 80 children [13]. The qXR (Qure.ai) is a regulatory-cleared CAD algorithm for tuberculosis screening and triage in individuals aged 15 years and above, with a well-established evidence base from multiple external validation studies [8]. The qXR-version 4.2.1 algorithm has recently undergone additional training specifically on paediatric dCXR images to interpret radiological features of tuberculosis in children under 15 years. However, its performance has not been independently validated in an exclusively paediatric population.

We therefore conducted an independent external validation to assess the diagnostic performance of the paediatric-optimized qXR algorithm for detecting pulmonary tuberculosis in children, using conventional reference standards (i.e., a microbiological reference standard [MRS] and a clinical reference standard [ClRS]) and Bayesian Latent Class Analysis (LCA), which statistically accounts for the absence of a perfect reference standard, an inherent limitation of tuberculosis diagnostic studies in children.

## Methods

### Study participants, settings, study design

We conducted a retrospective diagnostic accuracy study of the paediatric-optimized qXR algorithm using a repository of dCXR images from a well-characterised cohort of 932 Gambian children aged under 15 years with presumed pulmonary tuberculosis. Study participants were consecutively recruited through a childhood tuberculosis research programme at the Medical Research Council Unit The Gambia at London School of Hygiene & Tropical Medicine (MRCG at LSHTM), located in the Greater Banjul Area – a mixed urban, peri-urban, and rural region that includes the country’s capital city of Banjul. Recruitment was carried out across four prospective studies with identical protocols, evaluating preventive, screening, and diagnostic approaches for tuberculosis in children: (i) the Childhood Tuberculosis Programme Grant (February 2012–June 2017); (ii) Reach4Kids Africa (August 2017–December 2019); (iii) Reach4Kids Africa-2 (November 2021–October 2022); and (iv) Childhood Tuberculosis Research West Africa-2 (December 2022–November 2024). A detailed description of the aims, settings, screening, and recruitment procedures for these cohorts has been published previously [14–18].

Children with presumed pulmonary tuberculosis were enrolled through two pathways: active household contact tracing among those exposed to adults with newly diagnosed infectious tuberculosis disease, and clinical referral for diagnostic evaluation. At enrolment, participants underwent clinical assessment by a study paediatrician, a dCXR taken with a CARESTREAM DRX-Ascend system (combined with a Cannon CXDI 710CW detector; resolution 2800×3408 pixels), and sputum collection for microbiological testing. Sputum samples were obtained spontaneously or by induction using nebulised hypertonic saline, and were processed for pathogen detection by Xpert MTB/RIF or Ultra assay (Cepheid, Sunnyvale, USA), liquid culture in mycobacteria growth indicator tube (MGIT; Becton Dickinson, Franklin Lakes, NJ, USA), and solid culture on Löwenstein-Jensen medium. Positive cultures were confirmed as *Mycobacterium tuberculosis* (*Mtb*) by acid-fast staining and MPT64 antigen detection (Abbott, Palatine, IL, USA) or MTBDRplus line probe assays (Hain Lifesciences, Nehren, Germany).

### Tuberculosis classification

Using a combination of clinical, radiological, and laboratory findings, children were classified into three categories based on the case definitions for classification of intrathoracic tuberculosis in children: (i) confirmed tuberculosis; (ii) unconfirmed tuberculosis; and (iii) unlikely tuberculosis (Table 1) [19]. All children with confirmed or unconfirmed tuberculosis were referred to dedicated Gambia National Leprosy and Tuberculosis Control Programme (NLTBCP) treatment centres and attended follow-up visits at two months and at end of treatment.

**Table 1:** Revised case definitions for classification of intrathoracic tuberculosis for diagnostic evaluation studies in children.

| Case definition | Criteria |
| --- | --- |
| **Confirmed tuberculosis** | Bacteriological confirmation of *Mtb* (culture and/or Xpert MTB/RIF or Ultra assay) from at least one respiratory specimen |
| **Unconfirmed tuberculosis** | Bacteriological confirmation NOT obtained AND at least two of the following: |
|  | • Symptoms/signs suggestive of tuberculosis |
|  | • Chest radiograph consistent with tuberculosis |
|  | • Close tuberculosis exposure |
|  | • Positive response to tuberculosis treatment (requires a documented positive clinical response to TB treatment – no time duration specified) |
| **Unlikely tuberculosis** | Bacteriological confirmation NOT obtained AND Criteria for "unconfirmed tuberculosis" NOT met |

Abbreviation: *Mtb*, *Mycobacterium tuberculosis*

### Ethics Statement

Ethical approval was obtained from The Gambia Government/MRC Gambia Joint Ethics Committee for this secondary analysis and the four contributing studies. Written informed consent was obtained from each participant’s parent or legal guardian, with assent from children aged 12 years and older, at the time of enrolment. Although all children enrolled in the four contributing studies were eligible for this retrospective analysis, inclusion was restricted to those with a dCXR taken at enrolment who consented to use of their data for future ethically-approved studies.

### Digital chest radiograph analysis

Digital chest radiographs were identified using study-specific identifiers and extracted in DICOM format from the MRCG at LSHTM Picture Archiving and Communication System (PACS). Participants’ embedded metadata were anonymised and transferred to a dedicated digital library. The dCXRs were analysed using an offline deployment of the qXR algorithm hosted on a high-performance personal computer; DICOM files were imported directly from the digital library. Following automated analysis, numerical abnormality scores ranging from 0.00 to 1.00 were generated for each radiograph. All qXR analyses were performed by study personnel blinded to participants’ clinical data and tuberculosis classifications.

An independent expert paediatric radiologist (the human reader), blinded to clinical data, tuberculosis classifications, and qXR scores, assessed all dCXR images. Radiograph quality was graded as satisfactory, average, or poor based on three technical criteria: depth of inhalation, rotational alignment, and beam penetration adequacy. Each dCXR was categorised as (i) normal, (ii) abnormal–unlikely tuberculosis, (iii) abnormal–likely tuberculosis, or (iv) unreadable, based on the presence or absence of radiological features suggestive of tuberculosis according to the Union Diagnostic CXR Atlas for tuberculosis in children [20]. Corresponding data on demographics, clinical features, laboratory results, and tuberculosis classification were extracted from individual study databases and merged for analysis.

### Statistical analysis

Given the secondary, retrospective nature of this study, no formal sample size calculation was performed. We evaluated the diagnostic performance of the qXR algorithm against the MRS and ClRS using the manufacturer-recommended threshold (score ≥0·50). Under the MRS framework, children with confirmed tuberculosis were defined as MRS-positive, while children with unconfirmed or unlikely tuberculosis were defined as MRS-negative. Conversely, the ClRS framework classified children with unconfirmed tuberculosis as ClRS-positive and those with unlikely tuberculosis as ClRS-negative. Performance was quantified using area under the receiver operating characteristic curve (AUROC) and point estimates of sensitivity and specificity, each reported with 95% confidence intervals (CI). Performance was also assessed in pre-specified subgroups: children under five years versus those aged five to 14 years; children with Body Mass Index-for-age Z-score (BAZ) <-2 versus those with BAZ ≥-2; children with HIV (CHIV) versus HIV-uninfected children; and actively traced tuberculosis contacts versus children referred for diagnostic evaluation.

As all four tests evaluated (i.e., GeneXpert MTB/RIF/Ultra [Xpert], mycobacterial culture, qXR-version 4.2.1, and expert paediatric radiologist assessment) are imperfect, with sensitivities and/or specificities below 100%, we applied Bayesian LCA to estimate the sensitivity and specificity of each test [21]. This approach models the probability of true tuberculosis status as a latent variable, accounting for the covariance structure between tests. We first assumed conditional independence among all four tests, then relaxed this assumption by modelling conditional dependence between qXR and the human reader (both based on dCXR reading), and between Xpert and culture (both performed using the same sputum specimen). Full details of the Bayesian LCA modelling strategy, including prior specifications and model convergence assessments, are provided in the **Supplementary Material**.

Youden’s index analysis was performed to identify the qXR abnormality score threshold yielding the maximum combined sensitivity and specificity. Receiver operating characteristic (ROC) analysis was used to determine study-specific qXR analytical threshold scores, verified against both MRS and ClRS, at the WHO-endorsed minimum target product profile (TPP) sensitivity of 90% and specificity of 70% for a tuberculosis triage test [22].

All analyses were performed using Stata/SE version 17.0 (StataCorp, College Station, Texas, USA) and the *runjags* package in R for Bayesian modelling. This study is reported in accordance with the Standards for Reporting of Diagnostic Accuracy Studies (STARD) guidelines [23].

## Result

### Study population

Between 1 February 2012 and 31 October 2024, 2484 children with presumed pulmonary tuberculosis were enrolled across the four contributing studies. Of these, 1552 were excluded from the main analysis: 819 had no dCXR available, 729 were household contacts classified as having latent tuberculosis based on tuberculin skin test results and symptom screening, and four had a qXR read error. The final analysis included dCXR images of 932 children (**Figure 1**).

**Figure 1:**
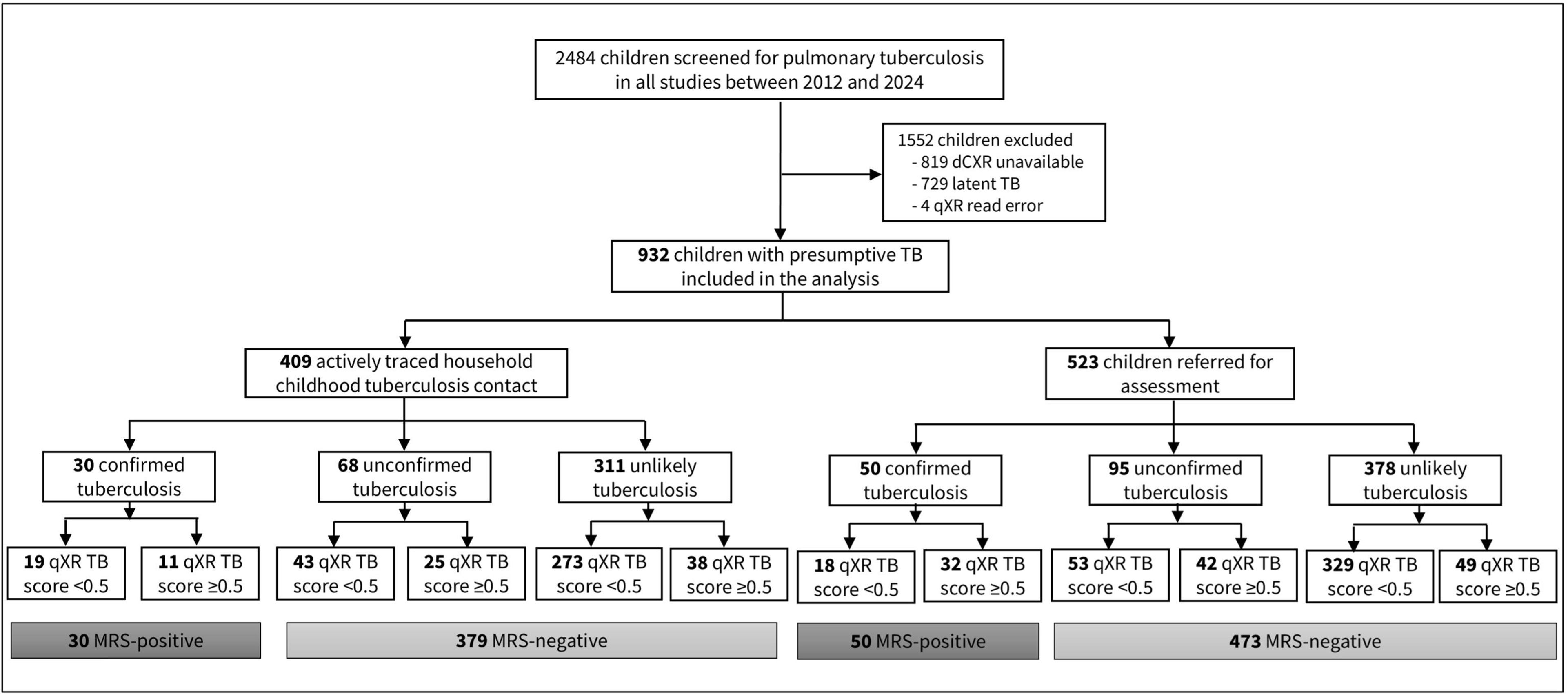
STARD diagram reporting the flow of participants in the study. Abbreviation: dCXR, digital chest x-ray; TB, tuberculosis MRS, microbiological reference standard **Alt text:** STARD flow diagram illustrating enrolment of children with presumed tuberculosis, exclusions with reasons for exclusion, pathway of participant recruitment, microbiological results and classification by paediatric-optimized qXR algorithm based on the manufacturer recommended threshold score (qXR score ≥ 0.50).

In the study population, 80 (9%) children were classified as confirmed tuberculosis, 163 (17%) as unconfirmed tuberculosis, and 689 (74%) as unlikely tuberculosis. The median age of the study population was 5.0 years (IQR 2.4–8.4), with 446 (48%) children under five years of age, 432 (46%) females, 409 (44%) actively traced household tuberculosis contacts, and 46 (5%) children with HIV. Full demographic and clinical characteristics are presented in **Table 2**. The distribution of qXR TB abnormality scores by tuberculosis classification is shown in the **Supplementary Figures 1–2.**

**Table 2:**
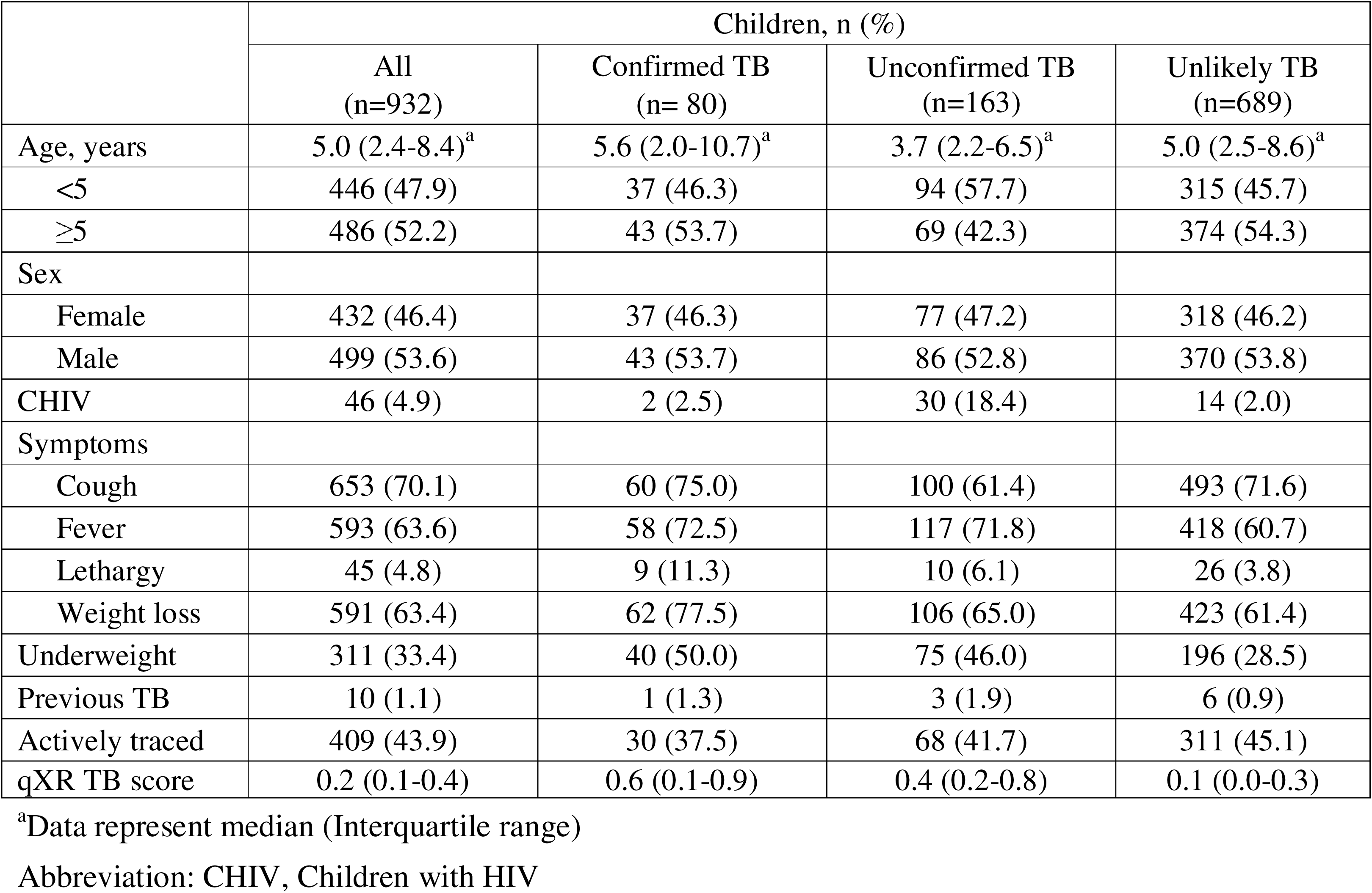
Demographic and clinical characteristics of participants included in the analysis.

### qXR performance against conventional reference standards

At the manufacturer-recommended threshold (score ≥0.50), qXR showed an AUROC of 0.68 (95% CI 0.61–0.75; **Figure 2a**), with a sensitivity of 54% (95% CI 43–64%) and specificity of 82% (95% CI 79–84%) against the MRS (**Figure 3a**). Against the ClRS, the AUROC was 0.73 (95% CI 0.69–0.77; **Figure 2b**), with a sensitivity of 41% (95% CI 34–49%) and specificity of 87% (95% CI 84–89%; **Figure 3b**).

**Figure 2a:**
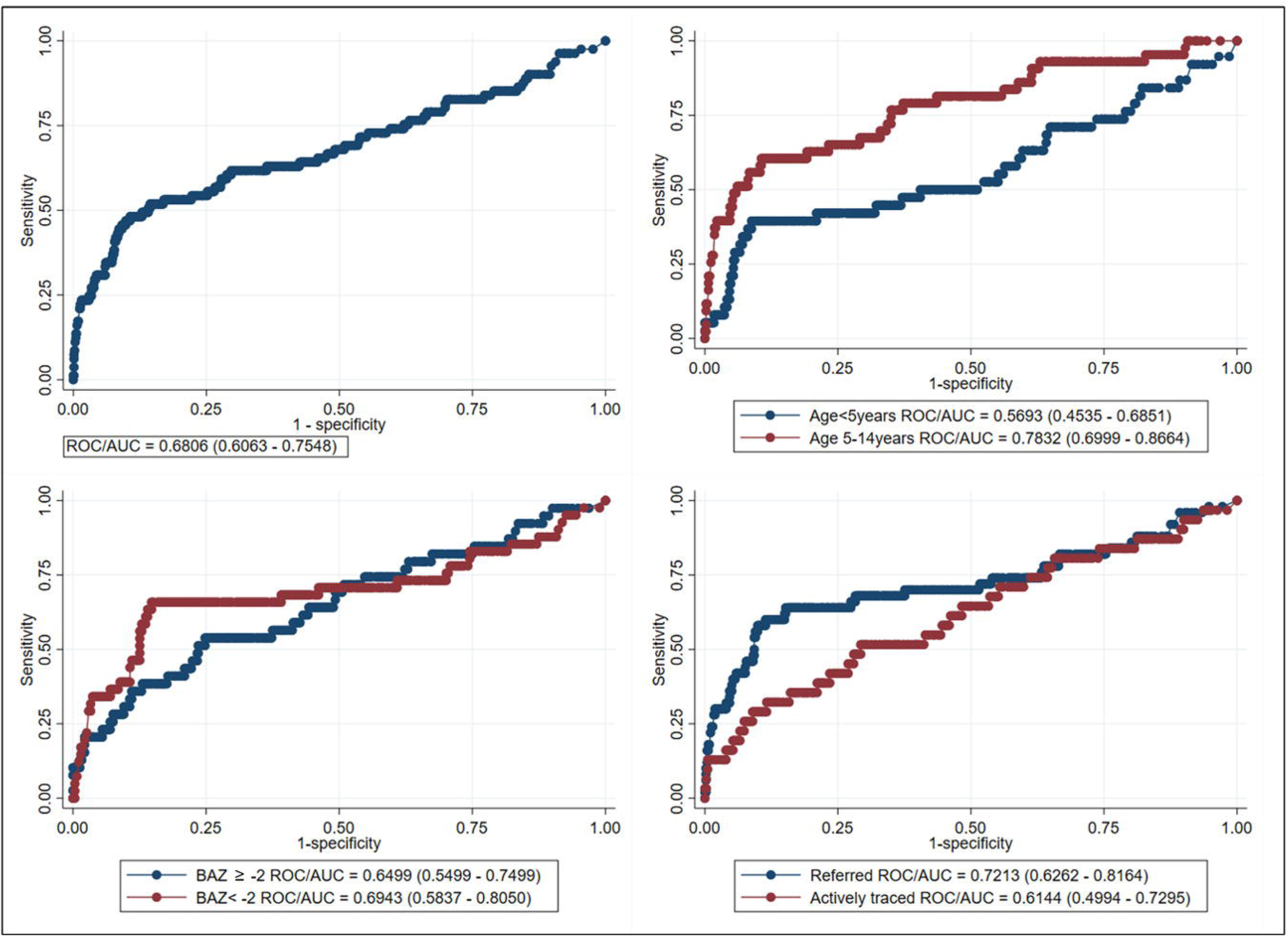
AUROC: (a) Total, (b) Age, (c) Nutritional status, and (d) Source (contact traced vs referred), against microbiological reference standard. Abbreviation: BAZ, Body Mass Index-for-Age Z-score **Alt text:** Receiver operating characteristic curves showing the diagnostic performance of the paediatric-optimized qXR algorithm in distinguishing microbiologically confirmed tuberculosis from unconfirmed and unlikely tuberculosis in all children in the study, children under five years versus those aged five to 14 years; children with Body Mass Index-for-age Z-score (BAZ) <-2 versus those with BAZ ≥-2; and actively traced tuberculosis contacts versus children referred for diagnostic evaluation.

**Figure 2b:**
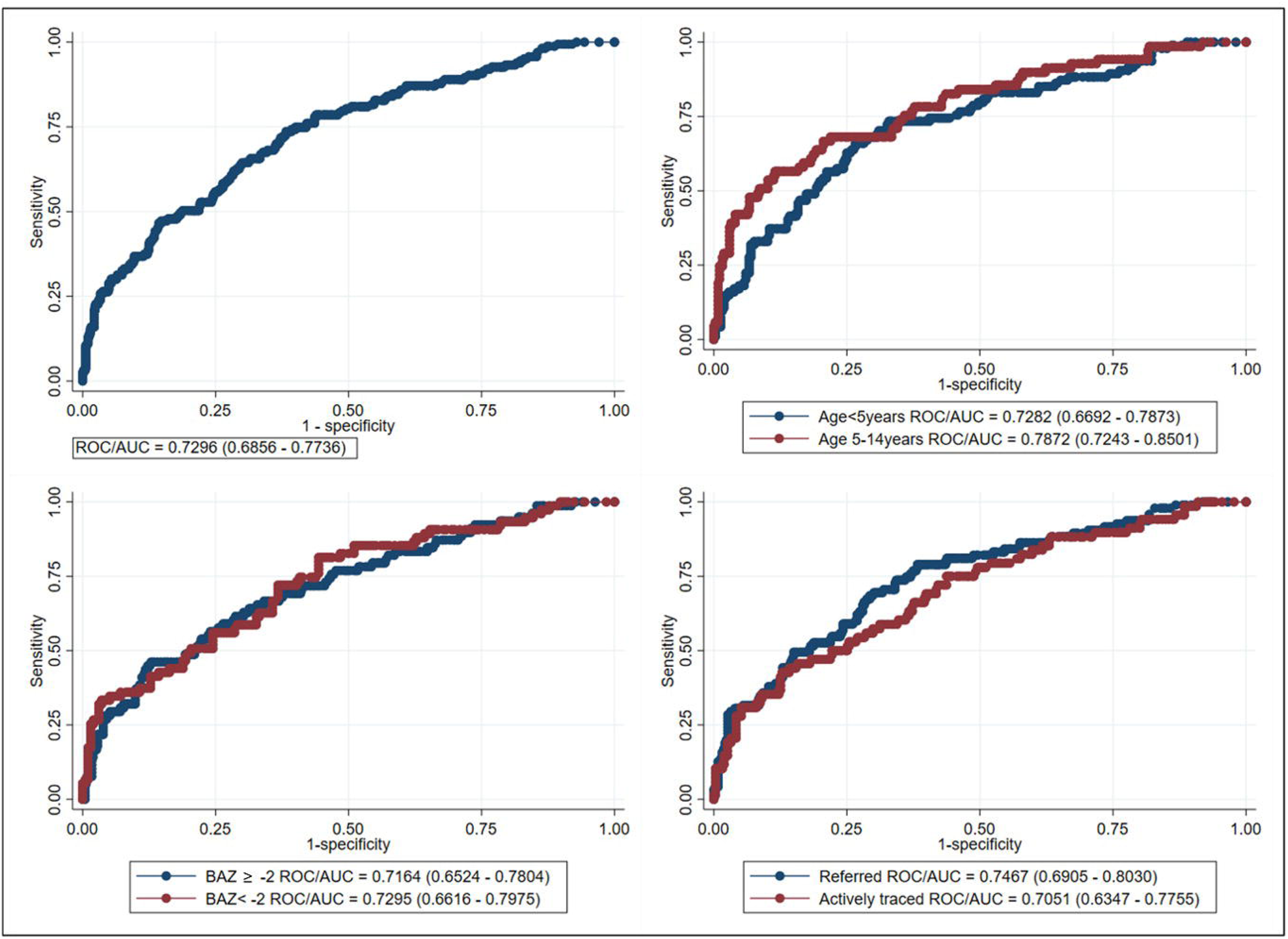
AUROC: (a) Total, (b) Age, (c) BAZ score, and (d) Source (contact traced vs referred), against clinical reference standard. Abbreviation: BAZ, Body Mass Index-for-Age Z-score **Alt text:** Receiver operating characteristic curves showing the diagnostic performance of the paediatric-optimized qXR algorithm in distinguishing unconfirmed tuberculosis (clinically and radiologically diagnosed tuberculosis) from unlikely tuberculosis in all children, children under five years versus those aged five to 14 years; children with Body Mass Index-for-age Z-score (BAZ) <-2 versus those with BAZ ≥-2; and actively traced tuberculosis contacts versus children referred for diagnostic evaluation.

**Figure 3a:**
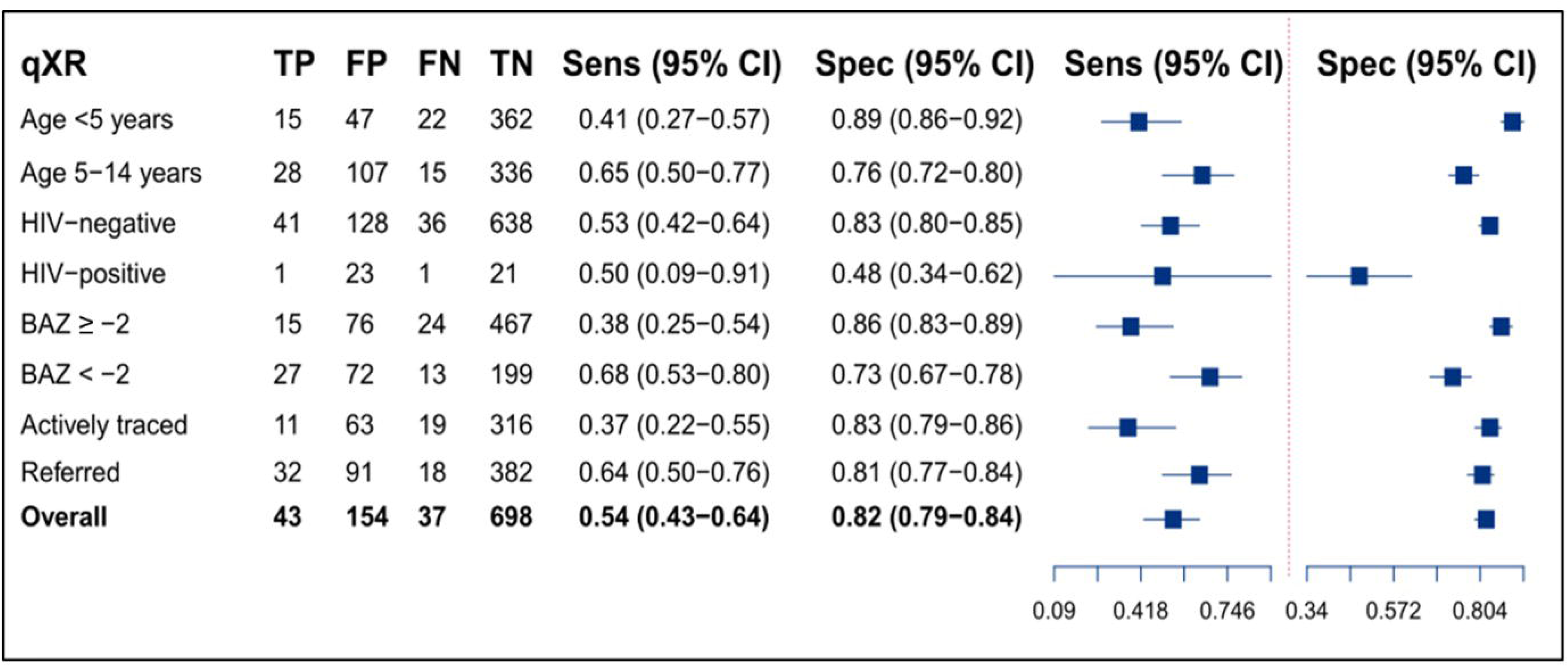
Sensitivity and specificity of qXR using the manufacturer-recommended threshold verified against microbiological reference standard. Abbreviation: TP, True Positive; FP, False Positive; FN False Negative; TN True Negative; Sens, Sensitivity; Spec, Specificity; BAZ, Body Mass Index-for-Age Z-score; CI, Confidence Interval **Alt text:** Forest plot showing the point estimates of sensitivity and specificity of the paediatric-optimized qXR algorithm in distinguishing microbiologically confirmed tuberculosis from unconfirmed and unlikely tuberculosis in all children, children under five years versus those aged five to 14 years; children with Body Mass Index-for-age Z-score (BAZ) <-2 versus those with BAZ ≥-2; children with HIV (CHIV) versus HIV-uninfected children; and actively traced tuberculosis contacts versus children referred for diagnostic evaluation.

**Figure 3b:**
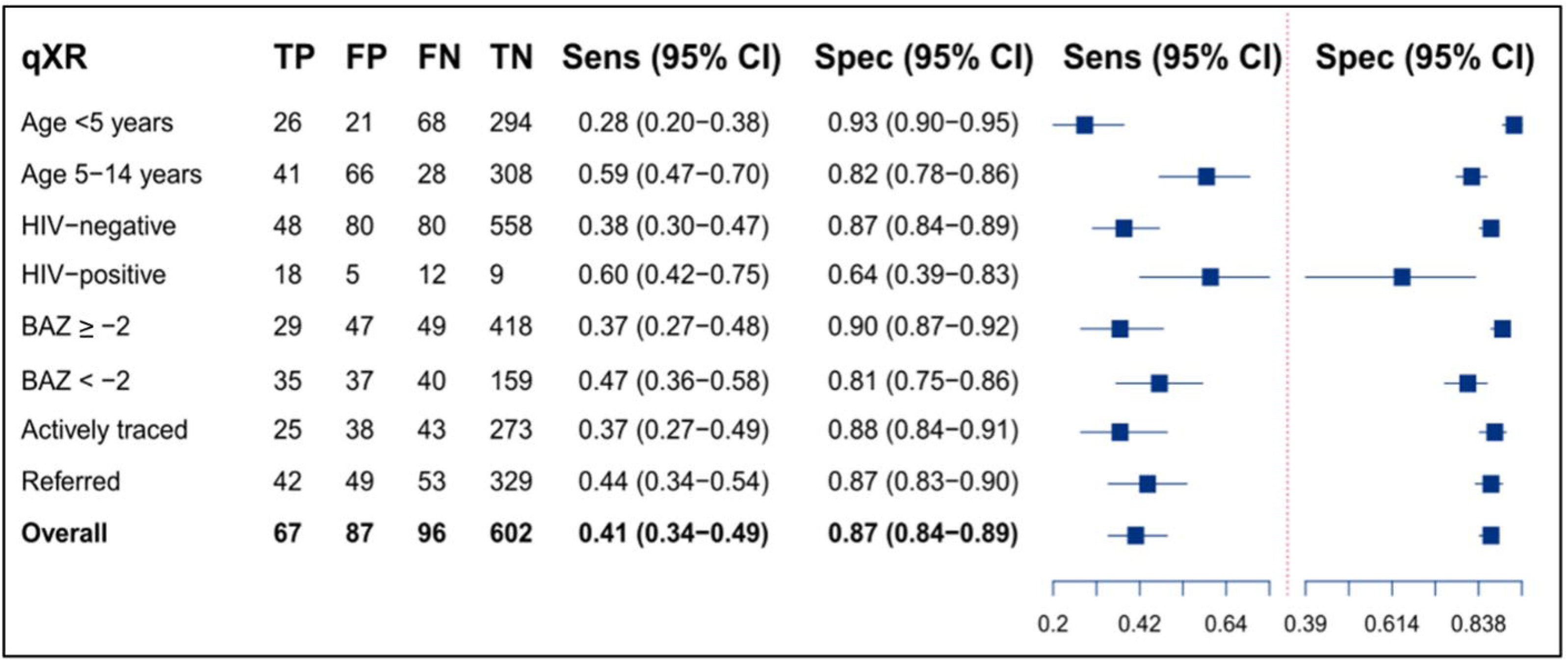
Sensitivity and specificity of qXR using the manufacturer-recommended threshold verified against clinical reference standard. Abbreviation: TP, True Positive; FP, False Positive; FN False Negative; TN True Negative; Sens, Sensitivity; Spec, Specificity; BAZ, Body Mass Index-for-Age Z-score; CI, Confidence Interval **Alt text:** Forest plot showing the point estimates of sensitivity and specificity of the paediatric-optimized qXR algorithm in distinguishing unconfirmed tuberculosis (clinically and radiologically diagnosed tuberculosis) from unlikely tuberculosis in all children, children under five years versus those aged five to 14 years; children with Body Mass Index-for-age Z-score (BAZ) <-2 versus those with BAZ ≥-2; children with HIV (CHIV) versus HIV-uninfected children; and actively traced tuberculosis contacts versus children referred for diagnostic evaluation.

Results of subgroup analysis are provided in the **Supplementary Material**. Notably, AUROC was lower in children under five years than in older children using both MRS (0.57, 95% CI 0.45–0.69 *vs* 0.78, 95% CI 0.70–0.87; **Supplementary Figure 3**) and ClRS (0.73, 95% CI 0.67–0.79 *vs* 0.79, 95% CI 0.72–0.85; **Supplementary Figure 5**). Similarly, AUROC was higher among children referred for clinical evaluation than among actively traced contacts under MRS (0.72, 95% CI 0.63–0.82 *vs* 0.61, 95% CI 0.50–0.73; **Supplementary Figure 3**) and ClRS (0.75, 95% CI 0.69–0.80 *vs* 0.71, 95% CI 0.63–0.78; **Supplementary Figure 5**). However, the confidence intervals overlapped across all subgroup comparisons.

### Head-to-head comparison of qXR with the human reader

Of the 932 dCXR images, 35 (3.8%) were classified as unreadable by the human reader, leaving 897 (96.2%) dCXRs available for head-to-head comparison. Against MRS, qXR demonstrated significantly higher overall discriminatory ability than the human reader (AUROC 0.65, 95% CI 0.59–0.71 *vs*. 0.54, 95% CI 0.49–0.60; p<0.001; **Figure 4**). At the manufacturer-recommended threshold, qXR demonstrated a sensitivity of 49.4% (95% CI 37.9–60.9%) and specificity of 80.9% (95% CI 78.1–83.6%), while the human reader had higher sensitivity (73.8%, 95% CI 62.7–83.0%) but substantially lower specificity (35.4%, 95% CI 32.1–38.7%).

**Figure 4:**
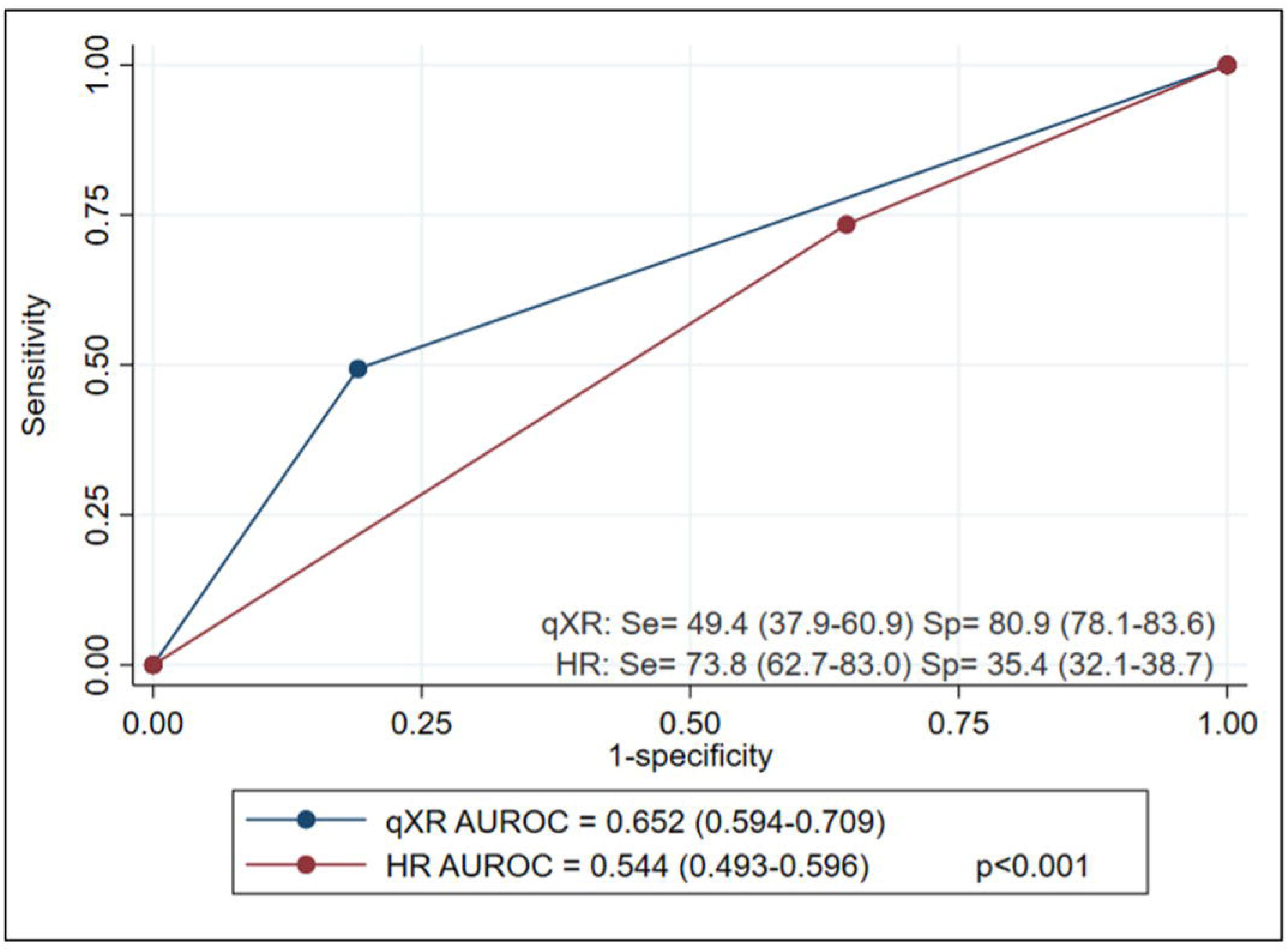
AUROC, sensitivity and specificity of qXR using the manufacturer-recommended threshold, and human reader verified against microbiological reference standard. Abbreviation: Se, Sensitivity; Sp, Specificity; HR, human reader; **Alt text:** Receiver operating characteristic curves showing the diagnostic performance of the paediatric-optimized qXR algorithm and the human reader in distinguishing microbiologically confirmed tuberculosis from unconfirmed and unlikely tuberculosis in all children.

### Bayesian Latent Class Analysis

Bayesian LCA was applied to the full study population and pre-specified subgroups to estimate test performance free from reference standard bias. Under the model assuming conditional dependence between qXR and the human reader, and between Xpert and culture, which was preferred on goodness-of-fit grounds, the estimated overall sensitivity and specificity of qXR were 88.6% (95% CrI 70.5–98.2%) and 81.8% (95% CrI 78.9–84.4%), respectively (**Table 3**). The corresponding sensitivity and specificity estimates for the human reader were 72.0% (95% CrI 57.5–83.7%) and 38.0% (95% CrI 34.7–41.3%), respectively. For Xpert, the sensitivity was 85.1% (95% CrI 67.2–95.7%) and specificity was 99.0% (95% CrI 98.0–99.6%), and culture showed sensitivity of 61.5% (95% CrI 41.8–80.4%) and specificity of 98.1% (95% CrI 96.9–99.1%). These results were consistent when allowing for conditional independence between all four tests in the model. The estimated specificities of qXR, Xpert and culture consistently exceeded that of the human reader overall and across all subgroups, except for CHIV. Full detail of the results of the Bayesian LCA are provided in the **Supplementary Material.**

**Table 3:** Estimated sensitivity and specificity of qXR, human reader, GeneXpert and culture as well as the prevalence of tuberculosis in study from Bayesian latent class analysis.

| Sample | qXR |  | Human reader |  | GeneXpert |  | Culture |  | Prevalence (95% CrI) |
| --- | --- | --- | --- | --- | --- | --- | --- | --- | --- |
|  | Sens (95% CrI) | Spec (95% CrI) | Sens (95% CrI) | Spec (95% CrI) | Sens (95% CrI) | Spec (95% CrI) | Sens (95% CrI) | Spec (95% CrI) |  |
|  | Assuming independence between all the four tests |  |  |  |  |  |  |  |  |
| Overall | 78.5 (64.8-89.3) | 81.5 (78.7-84.1) | 71.0 (59.4-80.9) | 38.1 (34.9-41.4) | 85.1 (69.4-95.4) | 98.9 (97.8-99.6) | 61.1 (46.5-75.2) | 97.5 (96.3-98.5) | 6.7 (5.0-8.8) |
| Referred | 82.5 (67.2-93.3) | 79.9 (76.0-83.5) | 68.0 (54.8-79.4) | 40.0 (35.7-44.5) | 84.6 (68.1-95.2) | 98.3 (96.5-99.3) | 64.0 (47.2-79.6) | 97.3 (95.5-98.6) | 8.7 (6.2-11.7) |
| Actively traced | 68.0 (42.9-88.3) | 82.9 (78.8-86.6) | 62.5 (45.5-77.3) | 40.7 (36.0-45.6) | 74.4 (49.9-92.0) | 98.3 (96.4-99.4) | 56.8 (33.5-80.0) | 96.9 (94.7-98.5) | 6.3 (3.9-9.7) |
| Age<5years | 69.8 (48.2-87.5) | 87.8 (84.1-90.9) | 68.5 (53.2-81.3) | 41.6 (36.9-46.4) | 78.3 (56.5-93.3) | 98.2 (96.3-99.4) | 59.7 (38.8-79.6) | 96.8 (94.6-98.3) | 7.3 (4.8-10.7) |
| Age≥ 5years | 83.6 (67.1-94.5) | 75.7 (71.5-79.6) | 63.8 (49.7-76.4) | 39.3 (35.0-43.7) | 83.3 (65.5-94.9) | 98.3 (96.6-99.4) | 62.7 (44.5-79.8) | 97.4 (95.6-98.7) | 7.9 (5.5-10.8) |
| BAZ ≥ -2 | 69.8 (48.1-87.6) | 85.4 (82.1-88.3) | 63.0 (47.4-76.8) | 42.6 (38.6-46.7) | 76.7 (54.6-92.5) | 98.6 (97.3-99.5) | 64.9 (43.6-84.0) | 96.9 (95.1-98.3) | 5.6 (3.7-8.2) |
| BAZ < -2 | 83.7 (67.2-94.6) | 72.1 (66.4-77.4) | 67.9 (53.9-80.0) | 36.9 (31.5-42.5) | 85.3 (68.3-95.7) | 97.1 (94.1-98.9) | 60.4 (42.1-78.5) | 97.2 (94.7-98.8) | 11.6 (8.1-15.8) |
| CHIV | 75.1 (35.5-96.5) | 52.0 (38.9-65.2) | 52.7 (32.9-71.8) | 53.4 (42.4-64.3) | 71.2 (42.9-91.3) | 92.1 (83.4-97.2) | 55.5 (24.0-85.5) | 92.9 (84.7-97.6) | 13.1 (7.2-21.4) |
| HIV-uninfected | 77.8 (63.8-89.0) | 82.9 (80.1-85.6) | 70.2 (58.3-80.4) | 39.3 (35.9-42.8) | 85.0 (69.4-95.3) | 98.8 (97.6-99.5) | 63.6 (48.7-77.6) | 97.3 (95.9-98.4) | 7.1 (5.3-9.4) |
|  | Assuming conditional dependence between qXR and HR, and GeneXpert and culture |  |  |  |  |  |  |  |  |
| Overall | 88.6 (70.5-98.2) | 81.8 (78.9-84.4) | 72.0 (57.5-83.7) | 38.0 (34.7-41.3) | 85.1 (67.2-95.7) | 99.0 (98.0-99.6) | 61.5 (41.8-80.4) | 98.1 (96.9-99.1) | 4.4 (2.9-6.3) |
| Referred | 89.8 (70.4-98.6) | 80.4 (76.4-84.0) | 68.4 (52.4-81.5) | 40.0 (35.6-44.5) | 84.3 (65.4-95.5) | 98.5 (96.8-99.4) | 65.9 (43.6-85.8) | 98.0 (96.2-99.2) | 5.8 (3.7-8.5) |
| Actively traced | 73.6 (37.6-95.8) | 83.3 (79.1-87.1) | 57.7 (38.1-75.2) | 40.7 (35.9-45.6) | 74.2 (47.3-92.3) | 98.4 (96.6-99.4) | 57.9 (28.2-85.6) | 97.5 (95.3-98.9) | 3.7 (2.1-6.2) |
| Age<5years | 76.3 (44.3-96.0) | 88.2 (84.5-91.5) | 62.9 (44.2-78.6) | 41.7 (36.9-46.6) | 77.8 (53.1-93.6) | 98.3 (96.5-99.4) | 61.6 (33.4-86.3) | 97.3 (95.2-98.8) | 4.4 (2.5-7.1) |
| Age≥ 5years | 89.5 (68.4-98.6) | 76.0 (71.8-80.0) | 64.4 (47.2-80.0) | 39.1 (34.7-43.6) | 82.5 (61.7-95.0) | 98.5 (96.9-99.5) | 63.6 (39.5-85.4) | 98.1 (96.3-99.3) | 5.1 (3.1-7.7) |
| BAZ ≥ -2 | 76.4 (43.7-96.1) | 85.8 (82.6-88.8) | 59.8 (40.9-76.3) | 42.6 (38.4-46.8) | 76.3 (50.7-93.0) | 98.8 (97.4-99.6) | 65.6 (36.7-88.8) | 97.4 (95.7-98.8) | 3.3 (1.9-5.3) |
| BAZ < -2 | 89.2 (68.1-98.5) | 72.5 (66.6-78.0) | 66.5 (49.5-80.5) | 36.8 (31.2-42.6) | 83.9 (64.0-95.5) | 97.3 (94.4-99.1) | 61.3 (37.3-84.0) | 97.7 (95.3-99.2) | 7.9 (4.8-11.9) |
| CHIV | 73.9 (33.6-96.3) | 53.3 (38.1-68.9) | 51.9 (32.1-71.0) | 57.5 (45.2-70.5) | 71.9 (43.9-91.7) | 91.4 (81.5-97.1) | 58.6 (26.3-87.4) | 91.8 (82.1-97.2) | 12.6 (6.7-21.2) |
| HIV-uninfected | 87.8 (68.7-98.1) | 83.3 (80.4-86.0) | 71.0 (56.2-83.0) | 39.2 (35.8-42.8) | 85.0 (67.1-95.7) | 98.9 (97.8-99.6) | 65.0 (44.6-83.8) | 97.9 (96.5-99.0) | 4.6 (3.0-6.6) |

Data are presented as %. Abbreviation: Sens, Sensitivity; Spec, Specificity; CrI, credible interval; BAZ, BMI-for-age z-score; CHIV, Children with HIV

### Threshold analysis

To assess the qXR threshold required to meet WHO-endorsed triage test criteria, we fixed sensitivity at the minimum TPP threshold of 90%: against MRS, this required a threshold of 0.019, yielding specificity of 14.4% (95% CI 12.1–17.0%); against ClRS, a threshold of 0.052 yielded specificity of 27.4% (95% CI 24.1–30.9%). Conversely, fixing specificity at the minimum of 70%: against MRS required a threshold of 0.345, yielding sensitivity of 61.7% (95% CI 50.3–72.3%); against ClRS, a threshold of 0.285 yielded sensitivity of 64.4% (95% CI 56.6–71.7%; **Table 4**).

**Table 4:** Accuracy estimates and qXR TB threshold score at WHO TPP targets for a triage test.

| **WHO TPP minimum requirement** | **Reference Standard** | **Sensitivity (95% CI)** | **Specificity (95% CI)** | **qXR threshold score<sup>a</sup>** |
| --- | --- | --- | --- | --- |
| Sensitivity set at 90% | *MRS* | 90.1<br>(81.5 - 95.6) | 14.4<br>(12.1 - 17.0) | 0.019 |
| Specificity set at 70% | *MRS* | 61.7<br>(50.3 - 72.3) | 70.1<br>(66.9 - 73.2) | 0.345 |
| Sensitivity set at 90% | *CIRS* | 90.2<br>(84.5 - 94.3) | 27.4<br>(24.1 - 30.9) | 0.052 |
| Specificity set at 70% | *CIRS* | 64.4<br>(56.6 - 71.7) | 70.0<br>(66.4 - 73.4) | 0.285 |

Data presented as %
<sup>a</sup>Data represent estimated qXR abnormality threshold score
Abbreviation: WHO TPP, World Health Organization Target Product Profile; CI, Confidence Interval; *MRS*, *Microbiological Reference Standard*; *CIRS*, *Clinical Reference Standard*

Using Youden’s index, the threshold maximising the combined sensitivity and specificity against MRS was 0.652, with a sensitivity of 48.1% (95% CI 36.9–59.5%) and specificity of 89.3% (95% CI 87.1–91.3%). Against ClRS, the optimal Youden’s index threshold was 0.215, with sensitivity of 73.6% (95% CI 66.2–80.2%) and specificity of 61.7% (95% CI 58.0–65.4%; **Supplementary Figure 7**).

## Discussion

This is the first independent external validation of a paediatric-optimized version of the qXR algorithm for tuberculosis detection in children. Against two conventional reference standards, qXR demonstrated moderate overall discriminatory ability (AUROC 0.68 and 0.73) with relatively high specificity (82% and 87%) but modest sensitivity (54% and 41%) at the manufacturer-recommended threshold. However, when the imperfect nature of these reference standards was accounted for using Bayesian LCA, estimated sensitivity increased substantially to 89%, while specificity remained at 82%. These findings suggest that conventional reference standards meaningfully underestimate the true diagnostic performance of qXR in children, and support the potential of the paediatric-optimized qXR algorithm as a complementary tool in childhood tuberculosis diagnostic pathways.

The moderate AUROC values of qXR observed against conventional reference standards are consistent with prior independent evaluations of CAD systems in exclusively paediatric populations. Previous studies have reported comparable AUROC values (0.68–0.72) against MRS, alongside similarly high specificity and low sensitivity profiles [12, 13, 24]. This pattern of high specificity and low sensitivity observed for CAD systems against MRS mirrors what has been reported for expert radiologist interpretation of chest radiographs for paediatric tuberculosis when assessed against MRS [25]. This convergence likely reflects a fundamental limitation of the MRS itself: in paucibacillary paediatric tuberculosis disease, microbiological confirmation may be absent even in children with true radiologically apparent tuberculosis, thereby artificially depressing the apparent sensitivity of any radiological test [26].

The Bayesian LCA results reinforce this interpretation. The conditional dependence model, which accounts for the biological correlation between qXR and human reader, and between Xpert and culture, estimated qXR sensitivity of 89%, substantially higher than the 54% estimated against MRS. Such upward revision aligns with prior applications of Bayesian LCA in childhood pulmonary tuberculosis, which showed that conventional MRS systematically underestimate true diagnostic sensitivity in this paucibacillary population [27]. These findings have direct implications for how future evaluations of CAD tools and other diagnostic tests for tuberculosis in children should be designed. Conventional reference standards alone are insufficient; Bayesian LCA should be incorporated as a primary analytical framework.

An important finding of this study is that the qXR algorithm identified radiological features consistent with tuberculosis in 41% of children who were not bacteriologically confirmed. Although this incremental yield is modest in absolute terms, it has meaningful clinical implications in settings where experienced paediatric clinicians and radiologists are unavailable. Given that paediatric tuberculosis is inherently a clinical and radiological diagnosis in most cases, the capacity of a CAD algorithm to flag children whose bacteriological workup is negative, but who may nonetheless have true disease, may reduce the risk of missed diagnoses and delayed treatment initiation. This complementary role for qXR is further supported by the significantly higher discriminatory ability of qXR compared with the human reader, particularly in terms of specificity, which consistently exceeded 80% for qXR across all reference frameworks, compared with less than 40% for the human reader.

The performance variation across subgroups warrants attention. AUROC was notably lower in children under five years of age compared with older children, reflecting the well-characterised diagnostic challenge in this age group, where radiological features of tuberculosis are most atypical and overlap substantially with other respiratory conditions [10]. Performance was also lower in actively traced contacts than in children referred for diagnostic evaluation, likely reflecting differences in disease prevalence and clinical severity across these groups. The observed reduction in specificity among children with HIV, whose radiological appearances are more heterogeneous, further underscores the need for subgroup-specific threshold calibration [7]. These patterns of variable performance are consistent with the recognised problem of biased AI algorithm performance in underrepresented sub-populations, and support the recommendation that paediatric tuberculosis training datasets for CAD development should be geographically diverse and should capture the full spectrum of radiological presentations across age groups [7, 28].

The threshold analysis highlights a fundamental tension inherent to any single-threshold operating strategy for tuberculosis triage in children. At no fixed threshold could qXR simultaneously meet the WHO-endorsed minimum TPP targets of 90% sensitivity and 70% specificity in this population, and the optimal Youden threshold yielded further performance trade-offs depending on the reference standard utilised. These findings are consistent with WHO guidance that CAD threshold selection should be context-specific, informed by the local tuberculosis prevalence, available laboratory capacity, and programmatic objectives – whether triage (prioritising sensitivity) or rule-out (prioritising specificity) [8]. They further highlight that an adult-optimized threshold is unlikely to be optimal for children, and reinforce the need for paediatric-specific threshold evaluation in any implementation study.

This study has some limitations. First, a proportion of dCXRs obtained before deployment of PACS at the study site could not be retrieved and were therefore unavailable for analysis. However, the distribution of tuberculosis classification among children included in the analysis closely mirrors that of the individual source studies, and published data from comparable childhood tuberculosis settings, making selection bias unlikely. Second, the number of children with confirmed tuberculosis was relatively small, which is reflected in the wide confidence intervals for MRS-based analyses. This is not due to the study design but the inherent difficulty of microbiological confirmation in paediatric tuberculosis. Third, the study was conducted in a West African country with a low/concentrated HIV prevalence. The findings may therefore not be directly generalisable to high HIV-prevalence settings, where altered radiological presentations and immune suppression may affect CAD performance. Finally, although this is an external validation study, it is retrospective, requiring prospective validation in diverse settings will before deployment recommendations can be made.

In conclusion, the paediatric-optimized qXR algorithm demonstrated moderate discriminatory ability and high specificity against conventional reference standards, with performance estimates substantially upward-revised by Bayesian LCA. These findings support the potential utility of paediatric-specific CAD as a complementary tool within childhood tuberculosis diagnostic pathways, particularly where experienced radiological interpretation is unavailable. However, further optimisation using well-characterised, geographically diverse paediatric dCXR datasets covering the full spectrum of tuberculosis radiological presentations across the age spectrum is necessary before qXR can achieve consistently adequate performance for routine clinical use. Prospective multisite validation studies, incorporating Bayesian LCA as a primary analytical framework, are needed to inform the evidence base for implementation of CAD system for tuberculosis in children.

## Funding

This study was supported by a Qure.ai Industry Collaboration grant (VEX060 to TT). The childhood tuberculosis studies that generated the stored digital chest radiographs and data used in this study were funded by a British Medical Research Council programme grant (MR/K011944/1 to BK), a United Kingdom Research and Innovation-Global Challenges Research Fund grant (MR/P024270/1 to BK), and a European and Developing Countries Clinical Trials Partnership-2 grant to the West African Networks of Excellence for TB, AIDS, and Malaria (WANETAM) consortium (CAS2020NoE-3103).

## Data Availability

Individual participant data underlying the results reported in this article, after deidentification, will be shared following the article publication and on reasonable request to the corresponding author.

## Acknowledgements

We thank the children whose digital chest radiographs and clinical data were used in this study and their parents or legal guardians and families. We also thank the field, clinic, and laboratory teams who facilitated the recruitment of the children in the original contributing studies. We acknowledge the contribution and support of the Data Management Team at the MRC Unit The Gambia at the LSHTM.

## Author Contributions

TT conceptualised the study. VFE, SCA, EN, SAO, NM, OMA, and TT contributed to the study design, implementation, and data acquisition. EN, SAO, AKS, UE, BK, and TT contributed to the data generation in the original contributing studies. VFE, SCA, EN, NM, and TT analysed and interpreted the data in this study. VFE and SCA wrote the first draft of the manuscript with input from EN, UE, BK, and TT. All authors contributed to the revision of the manuscript and approved the final manuscript version for submission.

## Disclaimer

The content is solely the responsibility of the authors. Qure.ai had no role in the study design, data acquisition, data analysis, data interpretation, writing of the manuscript, or decision to submit manuscript for publication. The corresponding author had full access to all study data and held final responsibility for the decision to submit the manuscript for publication.

## Declaration of interests

Authors have declared no conflict of interest

## Notes

### Competing Interest Statement

The authors have declared no competing interest.

### Funding Statement

This study was supported by a Qure.ai Industry Collaboration grant (VEX060 to TT). The content is solely the responsibility of the authors. Qure.ai had no role in the study design, data acquisition, data analysis, data interpretation, writing of the manuscript, or decision to submit manuscript for publication. The corresponding author had full access to all study data and held final responsibility for the decision to post the manuscript.
The childhood tuberculosis studies that generated the stored digital chest radiographs and data used in this study were funded by a British Medical Research Council programme grant (MR/K011944/1 to BK), a United Kingdom Research and Innovation-Global Challenges Research Fund grant (MR/P024270/1 to BK), and a European and Developing Countries Clinical Trials Partnership-2 grant to the West African Networks of Excellence for TB, AIDS, and Malaria (WANETAM) consortium (CAS2020NoE-3103).

### Author Declarations

The Gambia Government/MRC Unit The Gambia Joint Ethics Committee gave ethical approval for this work.

